# Impact of dexamethasone on persistent symptoms of COVID-19: an observational study

**DOI:** 10.1101/2021.11.17.21266392

**Authors:** A Milne, S Maskell, C Sharp, FW Hamilton, DT Arnold

**Author notes:** Corresponding author: Dr David Arnold, Academic Respiratory Unit, Southmead Hospital, BS10 5NB.

## Abstract

**Introduction:** Dexamethasone has been shown to reduce mortality for patients hospitalised with acute COVID-19 pneumonia. However, a significant proportion of patients suffer persistent symptoms following COVID-19 and little is known about the longer-term impact of this intervention on symptom burden.

**Methods:** Patients initially hospitalised with COVID-19 were prospectively recruited to an observational study (April-August 2020) with follow-up at 8 months (Dec 2020-April 2021) post-admission. A review of ongoing symptoms using a standardised systems-based proforma was performed alongside health-related quality of life assessment. In the UK, patients with COVID-19 (requiring oxygen) only received dexamethasone following the pre-print of the RECOVERY trial (June 2020), or as part of randomisation to that trial, allowing for a comparison between patients treated and not treated with dexamethasone.

**Results:** Between April to August 2020, 198 patients were recruited to this observational study. 87 required oxygen and were followed up at 8-months, so were eligible for this analysis. Of these 39 received an inpatient course of dexamethasone (cases) and 48 did not (controls). The groups were well matched at baseline in terms of age, comorbidity and frailty score. Over two-thirds of patients reported at least 1 ongoing symptom at 8-month follow-up. Patients in the dexamethasone group reported fewer symptoms (n=73, 1.9 per patient) than the non-dexamethasone group (n=152, 3.2 per patient) (p = 0.01).

**Conclusions:** In conclusion, in this case-control observational study, patients who received oral dexamethasone for hospitalised COVID-19 were less likely to experience persistent symptoms at 8-month follow-up. These are reassuring results for physicians administering dexamethasone to this patient group.

## Introduction

SARS-CoV-2 causes a wide spectrum of disease with a proportion of patients requiring hospitalisation for progressive respiratory failure with a 20% mortality rate (1). Many survivors of the acute phase of infection suffer from prolonged symptoms which has been termed Long Covid or Post-acute Sequelae of COVID-19 (PASC) (2). A large randomised platform trial run in UK hospitals throughout the pandemic demonstrated that the steroid, dexamethasone, reduced mortality in hospitalised patients receiving oxygen (3). This result was pre-printed in June 2020 and rapidly became standard of care for all patients hospitalised with respiratory failure (4). Despite its widespread use, little is known about the impact of dexamethasone on prolonged symptoms after COVID-19.

In this prospective case-control study, we assessed the symptom burden and quality of life of patients 8 months post initial hospitalisation with COVID-19, comparing individuals pre-and post the initiation of dexamethasone as routine treatment.

## Methods

Patients admitted to a single UK hospital with COVID-19 pneumonia were prospectively recruited to an observational study (from March to October 2020).

Patients were identified from the DISCOVER study, a single-centre prospective study (Bristol, UK) recruiting consecutive patients (≥18yo) admitted with COVID-19, full details published previously (5). Ethics approval via South Yorkshire REC: 20/YH/0121. The inclusion criteria were a positive PCR result for SARS-CoV-2 or a clinico-radiological diagnosis of COVID-19 disease. For the purposes of this sub-study, patients with an oxygen requirement during their admission were identified and grouped according to whether dexamethasone was prescribed during their admission (either as part of standard care post-June 16^th^ 2020 or when randomised to dexamethasone in the RECOVERY trial). Duration of dexamethasone course was recorded alongside routine demographic and clinical information. Patients who had received a course of steroids in the 2 weeks prior to admission or were on long-term steroids were excluded. Full definitions of clinical variables are available with the initial paper (5).

All patients were followed up at 8-months post initial admission, either by phone or in a face-to-face clinic (according to patient choice). At follow up, patients completed a ‘Short Form 36’ (SF-36) quality of life questionnaire (6) with review of ongoing symptoms using a standardised systems-based proforma.

Categorical variables were presented as counts with percentages. All continuous data were non-parametric and therefore presented with medians and interquartile range (IQR), unless otherwise specified. Differences between patient groups were evaluated using Mann Whitney-U and Kruskal Wallis tests for continuous data and Fisher’s exact test or Chi-squared testing for categorical data. Statistical significance was taken as p ≤ 0.05. Data were analysed using R version 4.0.0 with the packages “tidyverse” and “gtsummary”.

## Results

Between April to August 2020, 198 were recruited to this observational study. From this total, 87 required oxygen and were followed up at 8-months, so were eligible for this analysis. Of these 39 received an inpatient course of 6mg of oral dexamethasone once daily (cases) and 48 did not (controls). Patients received dexamethasone for 10 days or the duration of their hospital stay, whichever was shorter (median of 7 days, IQR 4-9). The groups were similar at baseline in terms of age, comorbidity, frailty score, and requirement for ventilation during admission (both non-invasive and mechanical ventilation). However, there were more men in the no dexamethasone group (69% vs 56%), and a higher rate of prior chronic lung disease (29% vs 15%) in the no dexamethasone group.

**Table 1:** Demographics

| Characteristics | No Dexamethasone Group<br>(N = 48) | Dexamethasone Group<br>(N= 39) |
| --- | --- | --- |
| Age | 60 (51,68) | 60 (53,72) |
| Male | 33 (69%) | 22 (56%) |
| Female | 15 (31%) | 17 (44%) |
| Diabetes Type 1 | 1 (2.1%) | 1 (2.6%) |
| Diabetes Type 2 | 5 (10%) | 10 (26%) |
| Heart Disease | 7 (15%) | 10 (26%) |
| Chronic Lung Disease | 14 (29%) | 6 (15%) |
| Severe Kidney Impairment | 3 (6.2%) | 3 (7.9%) |
| Hypertension | 15 (31%) | 8 (27%) |
| Rockwood score | 3 (2-4) | 3 (2-4) |
| Baseline PCR Positive | 36 (75%) | 36 (92%) |
| Non-invasive ventilation | 8 (17%) | 8 (21%) |
| Requirement for ITU | 6 (12%) | 4 (10%) |
| Length of Hospital Stay | 5 (3- 8) | 7 (4-9) |
| Mean duration (days) of Inpatient Dexamethasone Therapy (SD) | N/A | 7 (4-9) |

*PCR- Polymerase Chain Reaction, ITU- Intensive Care Unit, N/A- not applicable.*

### Symptomatology

Across the two groups (n=87), 68% of patients (n=59) reported at least 1 ongoing symptom at 8-month follow-up. Fatigue (44%), breathlessness (38%) and insomnia (29%) were the most frequently reported.

Figure 1 depicts the prevalence of ongoing symptoms at 8 months in each of the two groups. A total of 225 symptoms were reported across the cohort. Patients in the dexamethasone group reported significantly fewer symptoms (n=73, 1.9 per patient) than the non-dexamethasone group (n=152, 3.2 per patient), p=0.01 (KW Chi-Square 6.2). The largest differences were seen in fatigue (52 vs 33%, p=0.07) and insomnia (39 vs 15%, p=0.01). Patients in the dexamethasone group were more likely to be asymptomatic at follow-up, although this did not meet statistical significance. (41% vs 25%, p=0.11).

**Figure 1:**
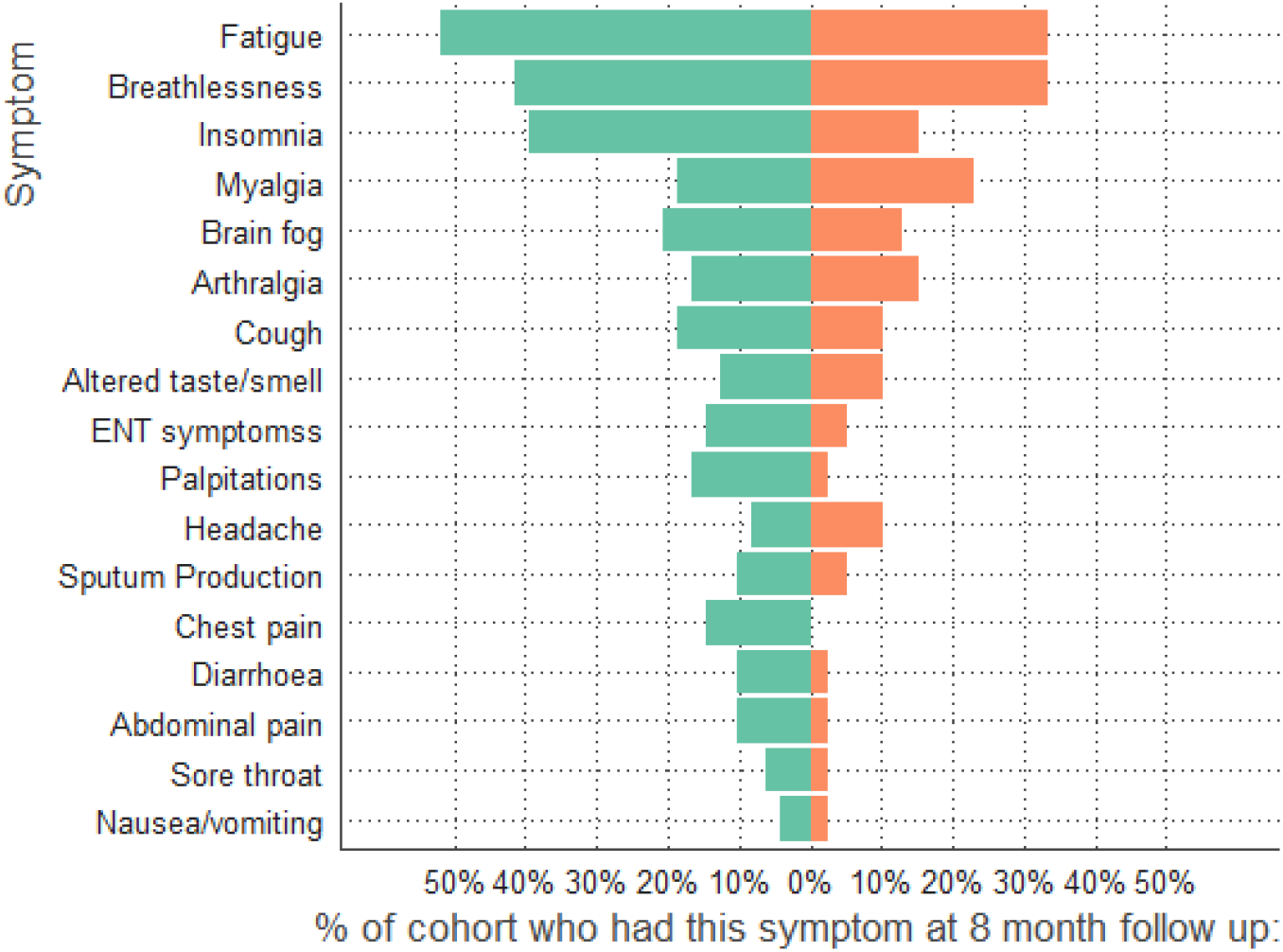
Symptoms at 8-month follow-up comparing dexamethasone group (orange) to no dexamethasone group (green)

### Mental and physical composite scores

The mental and physical composite scores (MCS and PCS) at 8 months were extracted from answers given to the SF-36 questionnaire by both groups. There was no significant difference in MCS or PCS between the groups with a median PCS of 43 (36-57) versus 49 (IQR 37-56) and median MCS of 33 (IQR 28-44) versus 37 (IQR 30-44) for dexamethasone and no dexamethasone groups respectively, p>0.1 for all comparisons.

## Discussion

Corticosteroids have been shown to improve short term outcomes in hospitalised patients with acute COVID-19 pneumonia. There is little evidence of their impact on longer term symptoms following resolution of the acute infection. This prospective case-control study demonstrates that symptom burden at 8-months was reduced in patients who were given a short course of dexamethasone in hospital.

SARS-CoV-2 infection causes a wide spectrum of disease in acute and post-acute infection. For patients hospitalised with acute COVID-19 pneumonia the RECOVERY trial has demonstrated that a course of the oral corticosteroid dexamethasone can significantly reduce 28-day mortality in moderate to severe cases. This led to a sea-change in the management of acute COVID-19 and has saved approximately 1 million lives worldwide (https://www.england.nhs.uk/2021/03/covidtreatment-developed-in-the-nhs-saves-a-million-lives/). However, steroids have an extended side effect profile even when given short-term. A sub-group analysis of the RECOVERY trial suggested that the benefit was restricted to patients requiring oxygen or already on a ventilator with those less severely affected actually harmed by a course of dexamethasone. Given the potent nature of steroids in this condition there are many outstanding questions that should be addressed with urgency including the optimum timing, dose and course (7). Another area of uncertainty that we have attempted to address is their impact of post-acute sequalae.

Following the acute phase of the disease a proportion of patients do not recover fully with persistent symptoms lasting for many months. This syndrome has been termed Long Covid or Post-acute Sequelae of COVID-19 (PASC) and although more likely to occur in the more severely affected in the acute phase is also common in patients who did not require hospitalisation (8). The syndrome is heterogenous in symptomatology, but the majority of hospitalised patients report fatigue, breathlessness, myalgia and insomnia. Two-thirds patients report ongoing symptoms at least 6 months after hospitalisation with COVID-19 (2). There are several hypotheses on the pathophysiology of Long Covid including persistence of virus, development of auto-immune disease or a product of organ damage during the acute phase. All might be theoretically affected by steroid use in either positive or negative direction. Reassuringly, this case-control study suggests that dexamethasone given in the acute phase of COVID-19 is associated with reduced long-term symptoms.

It is worth exploring potential reasons for this. Firstly, these results could represent confounding by indication, with patients receiving steroids more likely to recover regardless of the steroid prescription. However, in nearly all published data, patients with more severe disease have more prolonged symptoms after COVID-19, so we would expect the bias to favour more symptoms in those on dexamethasone. Secondly, dexamethasone use increased during the pandemic, after the RECOVERY trial results, and therefore most of the patients in the dexamethasone group were recruited later than those not on dexamethasone. Therefore, the reduced symptomatology could be due to general improvements in care that occurred during the pandemic which reduce long term symptoms. However, there is little evidence to support a reduction in long term symptoms of COVID-19 during the pandemic. Thirdly, the results could be due to patients with more severe disease dying and therefore being unable to be approached for follow up. This is by nature, difficult to account for, but as dexamethasone is known to improve mortality, we would expect the bias to artificially increase longer term symptoms in the dexamethasone group (by virtue of greater survival of a more frail population, who are known to have more symptoms after COVID-19). Finally, this association might be causal. Dexamethasone is known to impact on the severity of initial disease which is directly correlated with future symptom burden (8) and case-control studies of SARS-CoV-2 vaccination have shown that reducing severity of initial infection through vaccination reduces symptoms beyond 28 days (9).

There is currently little published research on the impact of steroid use on long term symptoms. An observational study remotely followed up hospitalised patients from the 1^st^ and 2^nd^ pandemic waves in Italy with a focus on taste and smell disturbance. They found that patients who received steroids were less likely to report ongoing chemosensory disturbance at 3 months (10). A multicentre observational study in the UK, recruited patients following hospital discharge with an assessment of symptoms and quality of life at 5 months post discharge. From 1077 patients discharged from 53 centres between March and November 2020, they found “no association between receiving systemic steroids during admission and recovery”, however, they did not limit the analysis for oxygen requirement during admission. Not doing so will heavily bias the assessment against steroids as, following the release of the dexamethasone results from the RECOVERY analysis in June 2020, the majority of patients on oxygen received steroids, prior to this date UK their use was restricted to the RECOVERY trial which included all hospitalised patients regardless of disease severity. By limiting our study to patients receiving oxygen during their admission we have reduced the impact of confounding by indication.

This study has weakness that reduce the generalisability of its findings. Firstly, it is a relatively small sample size, but this has allowed for detailed data to be collected with minimal data attrition. In addition, the analysis is limited to patients who required oxygen during their inpatient stay and are therefore quite severely compromised. The RECOVERY dexamethasone trial showed a trend towards harm in hospitalised patients not requiring oxygen and this analysis should not be used to justify steroid use in outpatients with acute COVID-19 or those with established persistent symptoms.

In conclusion, in this case-control observational study patients who received oral dexamethasone for hospitalised COVID-19 were less likely to experience persistent symptoms at 8-month follow-up. These will be reassuring results for physicians administering dexamethasone to this patient group.

## Data Availability

All data produced in the present study are available upon reasonable request to the authors, and with the limitations of confidentiality.

